# Making cotton face masks extra-protective by use of impervious cloth as the front layer to restrict flow of aerosols and droplets

**DOI:** 10.1101/2020.07.30.20165563

**Authors:** Priyanka L. Shahane-Kapse, M R Shende, Sumit Kar, Pradeep Deshmukh, Dhiraj Bhandari, Rahul Narang

## Abstract

**Introduction:** One of the important measures to prevent spread of COVID-19 in community is use of face mask. Though the debate is going on regarding the airborne transmission of SARS-CoV-2 it makes reasonable point for universal use of face masks. A large variety of face masks are available in the market or people can make their own using household items. The efficacy of masks depends upon the type of cloth and number of layers of the cloth.

**Material and methods:** We have created an innovative mask with two layers of cotton and an impervious layer. The impervious layer made from polypropylene coated with polyurethane was applied on the outer side in the middle half of the mask in front of mouth and nose. The efficacy of this test mask was measured against N95FFR (reference standard), triple layer surgical masks and single layer cotton mask. A manikin was used wearing these masks/respirator and aerosols/droplets of diluted red coloured carbol fuchsin and fluorescent Auramine O were sprayed from distance of 1m and 2m. We also tested use of face shield. Both macroscopic and microscopic examination of the dissected masks and respirator was performed.

**Results:** The N95FFR was able to block the aerosols/droplets by its front layer. One triple layer surgical mask showed microscopic presence of stain in its innermost layer while the other blocked it with middle layer. The single layer cotton mask was not able to protect as we observed stain on the face itself. The test mask blocked most of the stain on impervious layer and also on the front cotton layer on lateral sides, where impervious layer was absent. When fluorescent stain was used, ultraviolet examination demonstrated that the whole area covered by test mask was clean while the other non covered area was fluorescent.

**Conclusion:** We believe that our innovation can be used in the community as well as in general areas of the hospital like, offices, labs, etc. and can be a better alternative to single use triple layer surgical masks. Further testing may be done by other organizations to rule out bias in our study.

## Introduction

The airborne transmission of SARS-CoV-2 causing COVID-19 has been a debatable issue; however, the general agreement is that in closed spaces with poor ventilation and more human activity airborne transmission may occur.^1^ Considering this possibility, the universal use of face masks has been recommended even when social distancing is being followed.^2^ The face mask or a respirator in front of mouth and nose works by restricting the two way flow of droplets as well as aerosols. It has been proven that a person can shed SARS-CoV-2 while breathing, talking, sneezing and coughing by liberating viruses in varying numbers.^3^

The N95FFR has been recommended for hospital use while the general public has been recommended the use of face masks that are available in the market or can be prepared as do it yourself activity. (https://www.fda.gov/medical-devices/personal-protective-equipment-infection-control/n95-respirators-surgical-masks-and-face-masks) Guidelines for preparing homemade masks are available online.(https://www.cdc.gov/coronavirus/2019-ncov/prevent-getting-sick/how-to-make-cloth-face-covering.html) The cloth used for making such masks may vary in quality and a comparison of efficacy of different materials with respect to N95FFR has been performed by Lustig et al 2020.^4^

Triple layer surgical masks fall in category between N95FFR and general use face masks and are being used by personnel in general areas of the hospital.

In our previous study, we had used an impermeable cloth made up of polypropylene coated with polyurethane of 82 GSM to make re-usable gowns for hospital, laboratory and community.^5^ The cloth was tested for blood penetration test by South India Textile Research Agency (SITRA) and was approved as PPE material. It was further tested in our laboratory by repeated exposures to various disinfectants and heat. By now, we have tested the gowns made from same cloth by repeatedly exposing them to hypochlorite and autoclaving for at least 40 times and the integrity is still maintained. The cloth is light in weight and is very smooth as per feedback from end users. With the success of the material as impermeable cloth, we tested it further to create and test triple layer face masks.

## Material and methods

This study was conducted in the departments of Anatomy and Microbiology of the Mahatma Gandhi Institute of Medical Sciences, Sevagram, Wardha, India.

### Preparation of innovative masks

We prepared triple layer masks using two layers of cotton cloth and one outer layer of impervious cloth tested in our previous study.^5^ The three layers were made in the form of pleats as given in Photograph 1. The compressed mask is of 20cm in length and 8cm wide. After opening the pleats the width in the middle reaches 16cm. To make it impenetrable, we have used a third layer of impervious cloth in the middle 10cm area in front of nose and mouth. We left the lateral portions as double layer to permit flow of air. Four long strings were made on each side to tie them at the back of the head. (Photograph 1) The type of cotton used was similar to Kona cotton on microscopy.^4^ (Photograph 2)

**Photo 1.**
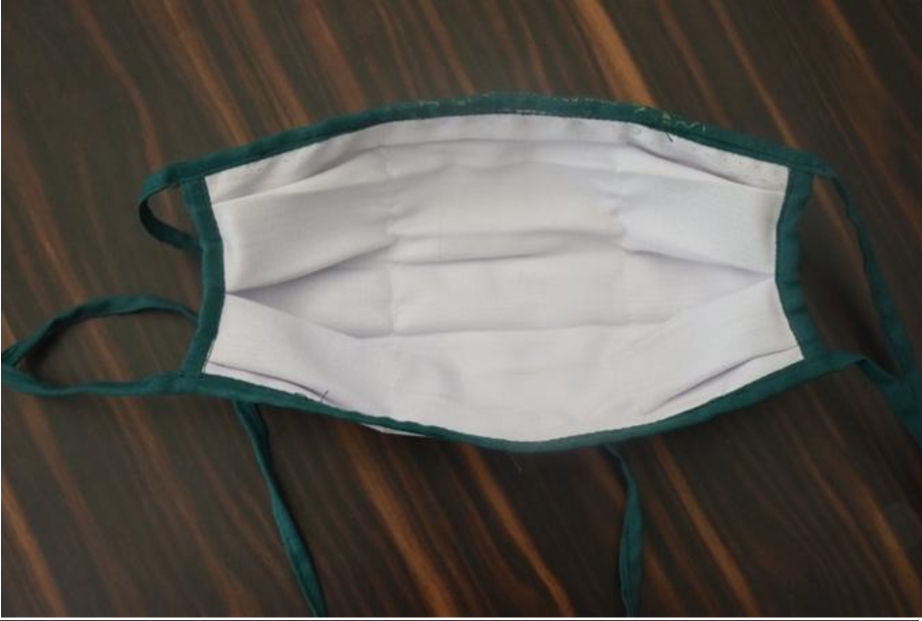
showing inside of the test mask with strings. (Length 18cm, width 8cm, width in the middle 16cm.)

**Photo 2:**
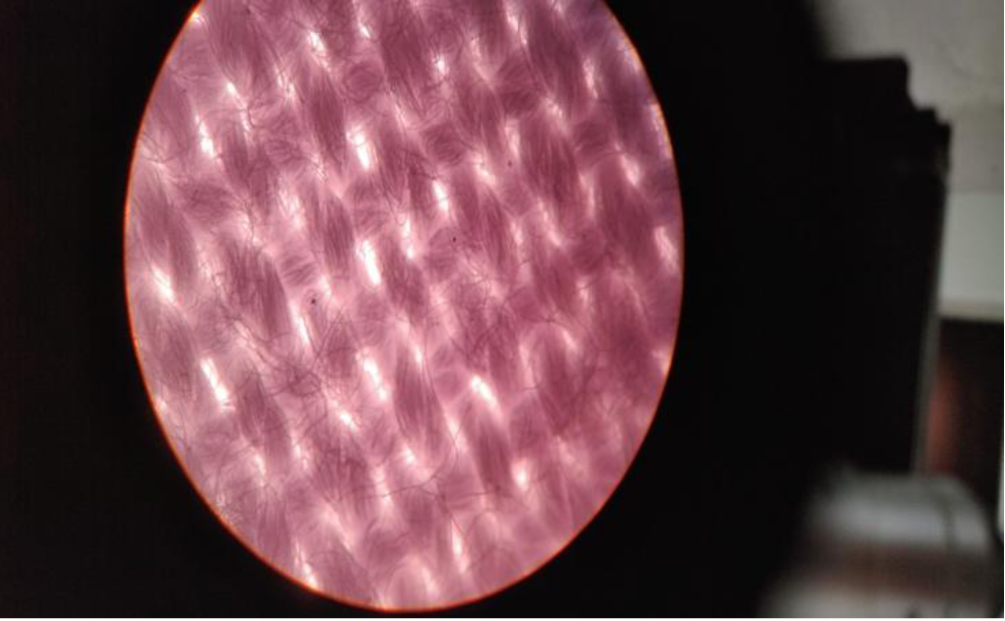
Inner layer of triple layer test mask created by our laboratory (50x)

This mask was tested along with single layer cotton mask, two varieties of triple layer surgical masks against N95FFR.

### Preparation of manikin

The testing was done on a manikin. We used an average size skeleton to recreate human face in the dissection hall of the anatomy department. The face was recreated using cotton and bandages. The lower part of the skeleton was covered with the same impermeable cloth to protect it. The face was exposed repeatedly to coloured aerosols and droplets. For this purpose, every time the face was covered with new cloth of the size 2×1feet. In front of the eyes laboratory goggles were used. The skeleton was hung using a hook. (Photograph 3)

**Photo 3.**
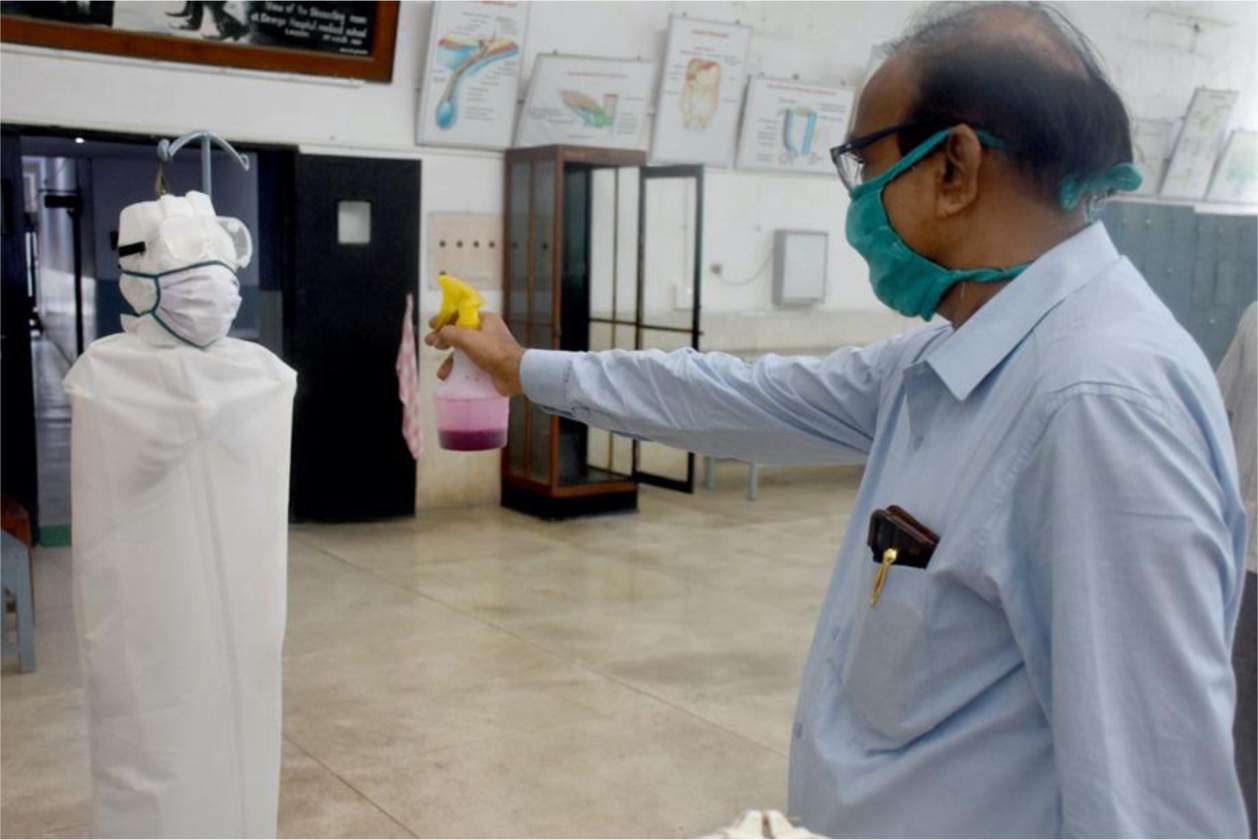
showing manikin, sprayer and anatomy dissection hall

### Liquid used to create droplets and aerosols

We used two varieties of coloured liquids-

i. For most of experiments - 1:10 solution of carbol fuchsin that is routinely used for staining in the laboratory.
ii. To look for the spread of aerosols/ droplets on face and clothes, we also tested spraying the manikin with fluorescent Auramine O dye. The post exposure examination was conducted using Wood’s Lamp with UV light of 365nm wavelength.

### Experiment

A spray bottle was used from a distance of 1m and 2m. (Photograph 3)

For all the experiments while manikin was wearing masks, a distance of 1m was used.

The manikin was exposed to dilute stain spray in the following manner-

1. Without any protection from 1m and 2m
2. With face shield from 1m and 2m
3. With face shield and masks from 1m and 2m
4. Without face shield from 1m using
  a. triple layer surgical mask number 1 (Green)
  b. triple layer surgical mask number 2 (Blue)
  c. single layer cotton mask
  d. triple layer mask under testing
  e. N95FFR (Reference Standard)

To further test the efficacy of the test mask, we also conducted another experiment where the mask wearing manikin was exposed to liquid spray containing Auramine O to check the spread of fluorescence on face and other parts of body.

All these experiments were photographed and for certain steps HD videos were also made.

### Examination of spread of aerosols and droplets on masks/respirators

After each experiment, gross examination of the manikin was done for spread of aerosols and droplets. The gross examination of the outer and inner surfaces of masks as well as respirator was also performed. The used masks and respirators were dissected and wherever the droplets were visible on outer layer, all the three layers was examined by microscopy using 50x and 100x of a binocular microscope. (Olympus CH20iBIMF)

We also studied convenience of the end users by interviewing 10 persons who had used the masks during last 10days. The cost was calculated from the price of cloth and stitching.

## Results

The crude velocity of the sprayed aerosols/droplets was calculated using high definition video and was approximately 180km/hr. We could not measure the size of droplets released from the sprayer.

### Without any protection from 1m and 2m

From 1m whole of the manikin was covered with spray with larger drops visible on lower side and fine drops on the face. While using a distance of 2m, we had to create air turbulence using a pedestal fan to carry aerosols to the manikin. (Photograph 4)

**Photo 4.**
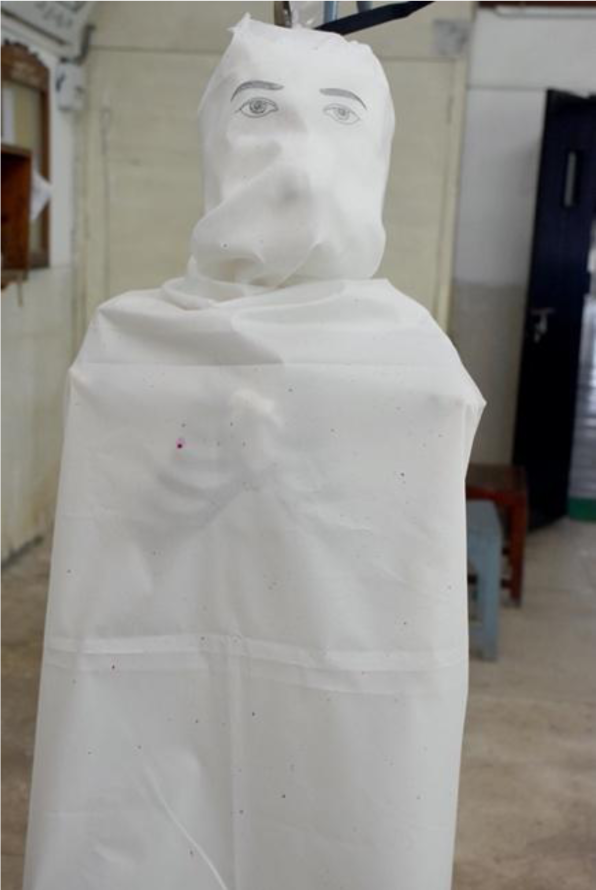
showing large droplets on body and fine droplets on face

### With face shield from 1m and 2m

The use of face shield protected the face from both the distances.

### With face shield and masks from 1m and 2m

There was no stain visible on the mask and face when face shield was used. (Photograph 5)

**Photo 5.**
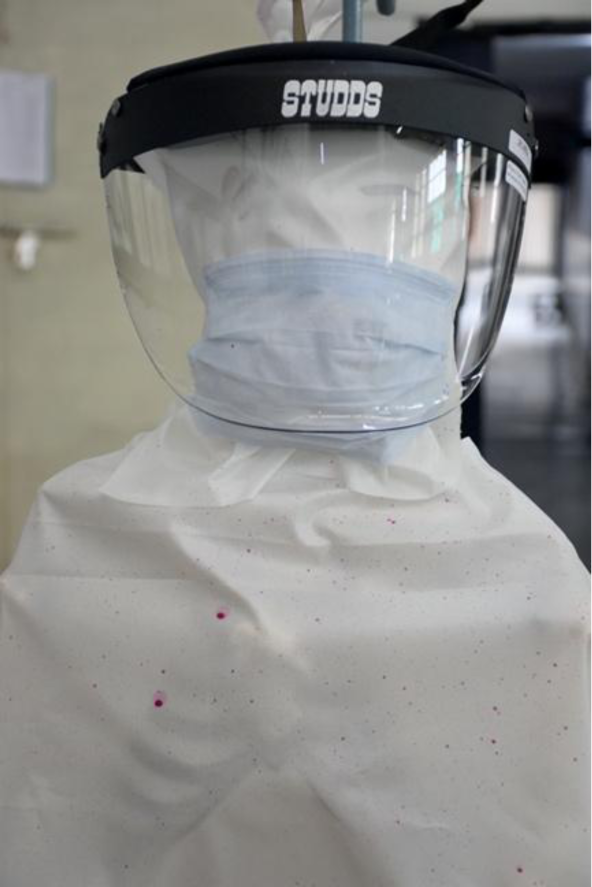
showing face shield and mask and stains on surface of shield and body

### Without face shield from 1m using various masks and respirator

a. Triple layer surgical mask number 1 (Green)- On gross examination, the stain was visible on front surface, while it was not visible on the back surface. However, when the dissection of three layers was carried out, we found one spot where the stain was visible on all the three layers on microscopic examination. (Photographs 6-9)

**Photo 6:**
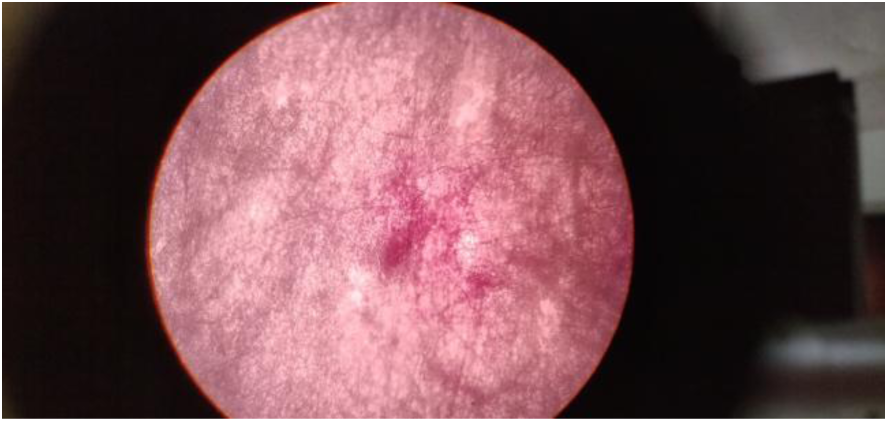
Stain observed in the outer layer of triple layer surgical mask (50x)

**Photo 7:**
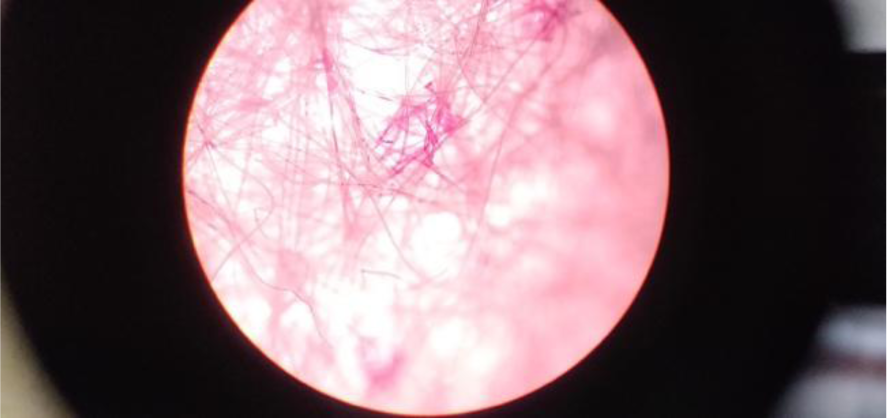
Stain observed in the middle layer of triple layer surgical mask (50x)

**Photo 8:**
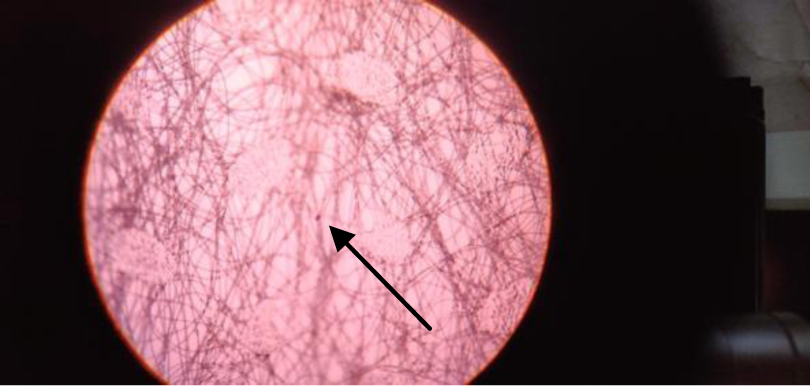
Stain observed in the inner layer of triple layer surgical mask (50x)

**Photo 9:**
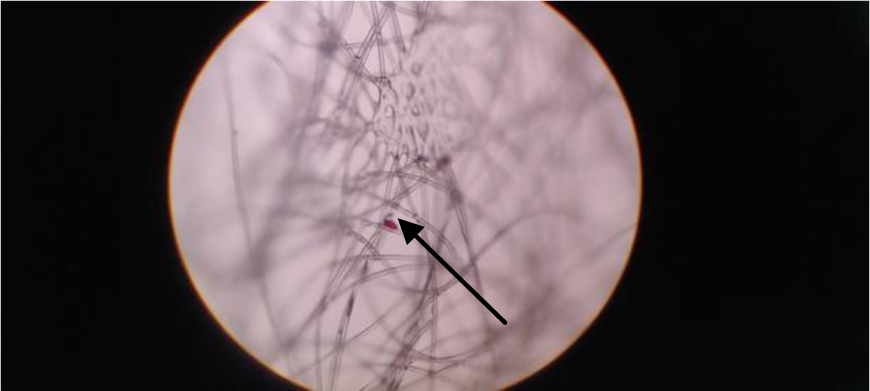
Stain observed in the inner layer of triple layer surgical mask (100x)
b. Triple layer surgical mask number 2 (Blue)- Here the front and middle layers had macroscopic and microscopic evidence of stain, while the innermost layer was clear both macro-as well as microscopically.
c. Single layer cotton mask-In this the stain was able to penetrate the single layer and reach the face of manikin and fine coloured spots were visible on the face.
d. Triple layer mask made by our laboratory (Test mask)- In the middle half where the impermeable cloth was there the stain remained on the outmost layer and did not penetrate it. On the lateral side, few spots were visible on white cotton. On macroscopic and microscopic examination, the stain was absent on the inner layer even when only two layers were present. (Photographs 10-11)

**Photo 10.**
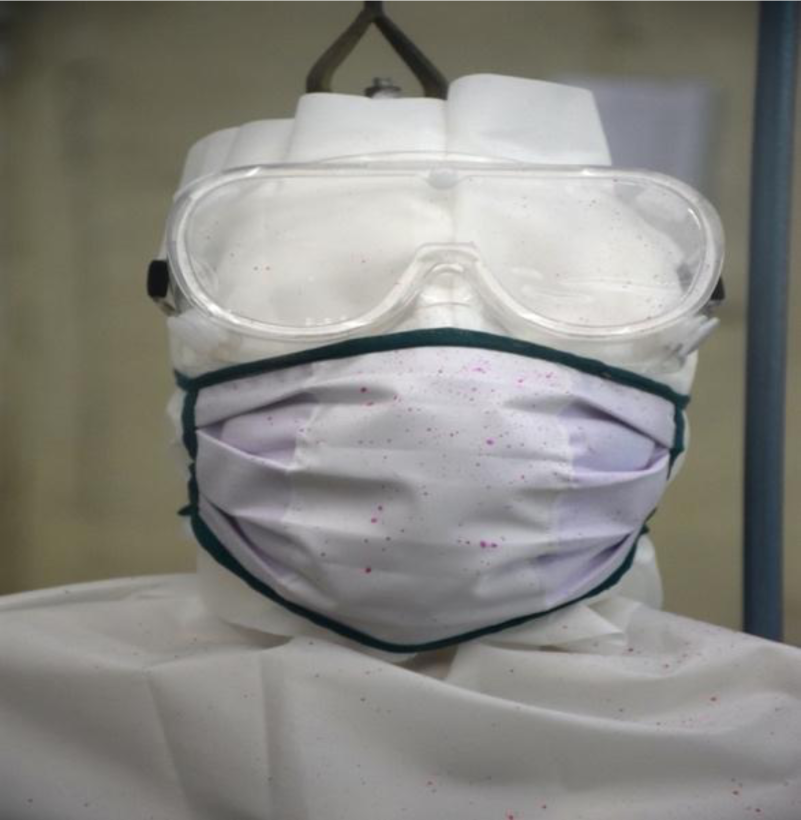
showing middle impervious layer of test mask stained with aerosols and droplets

**Photo 11:**
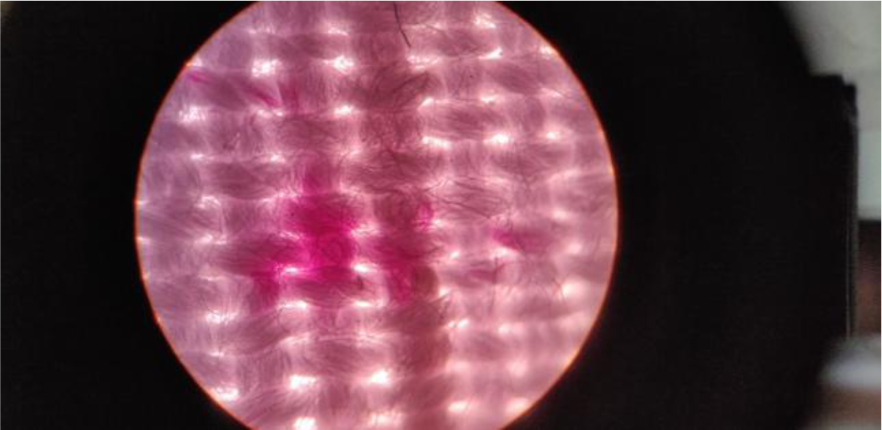
Stain observed in the outer layer of triple layer test mask on microscopy (50 x)
e. N95FFR: In this only outermost layer had evidence of stain on macro and microscopic examination.

**Table 1:**
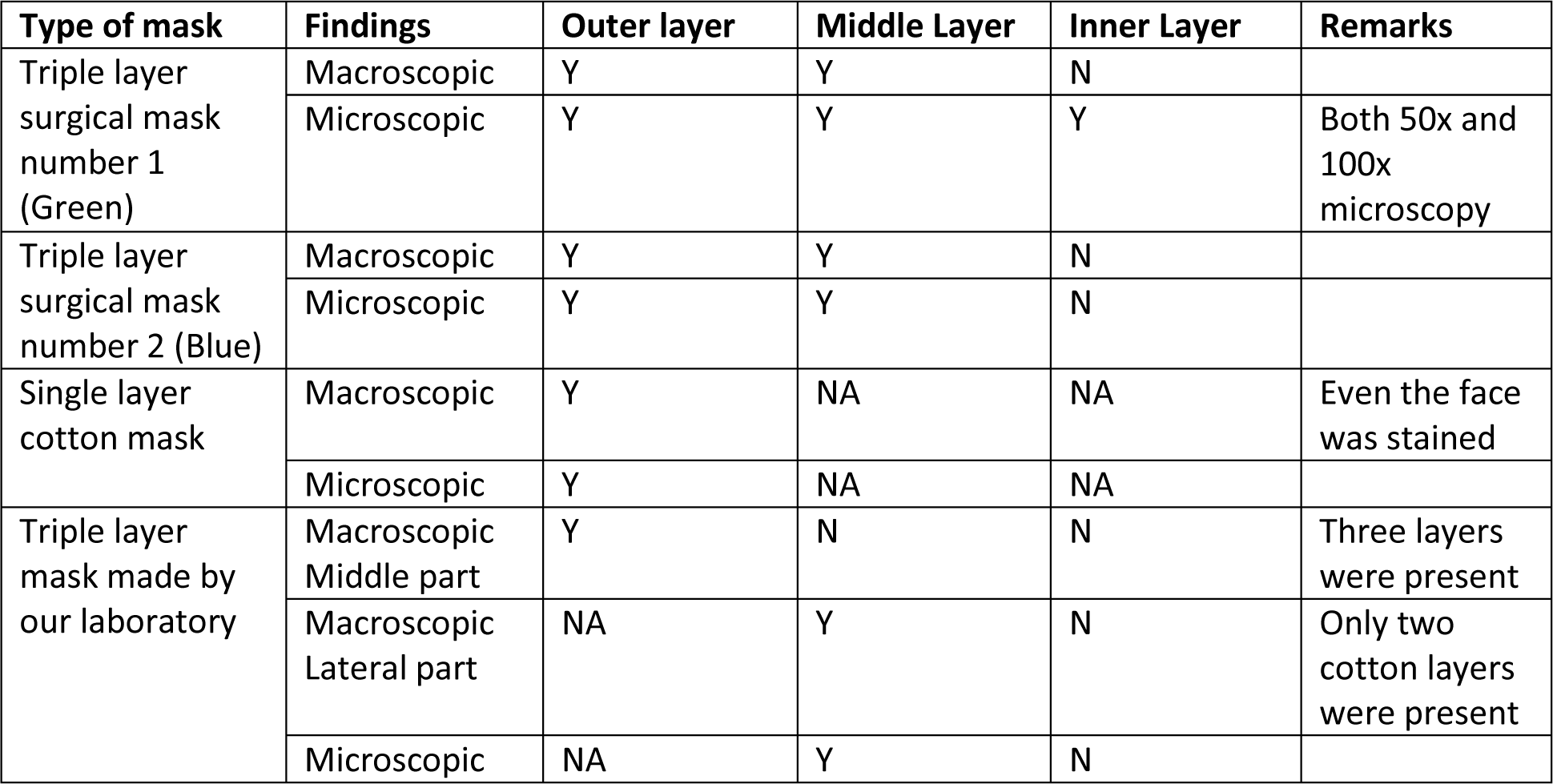
Comparison of macroscopic and microscopic findings on presence of aerosols/droplets on face mask

In the second experiment with test mask and fluorescent dye, it was observed that the whole of front of the mask as well as exposed parts of the face and body were fluorescing indicating the wide spread of aerosols. The mask was then removed to check if stain had penetrated the mask. No fluorescence was observed on the area covered by the mask. (Photographs 12-14)

**Photo 12.**
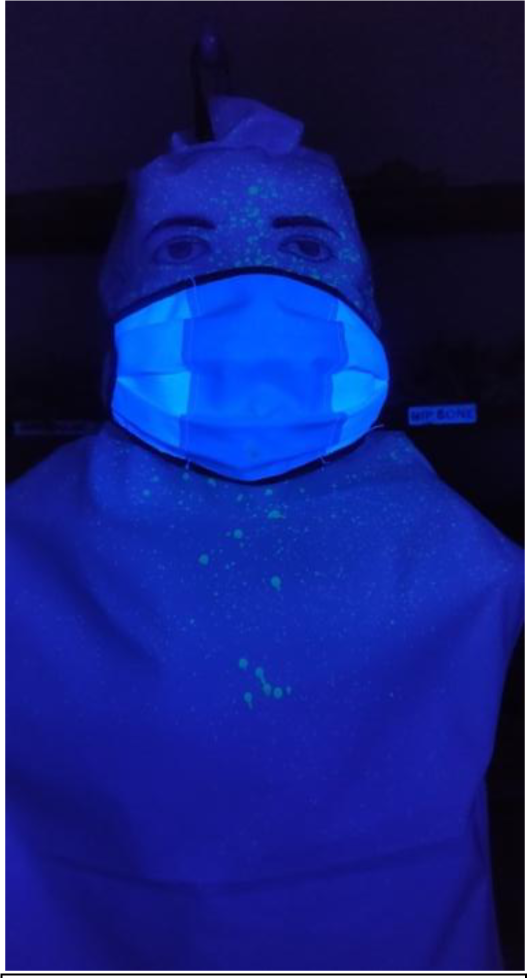
showing spread of aerosols/droplets on face and body under UV light

**Photo 13.**
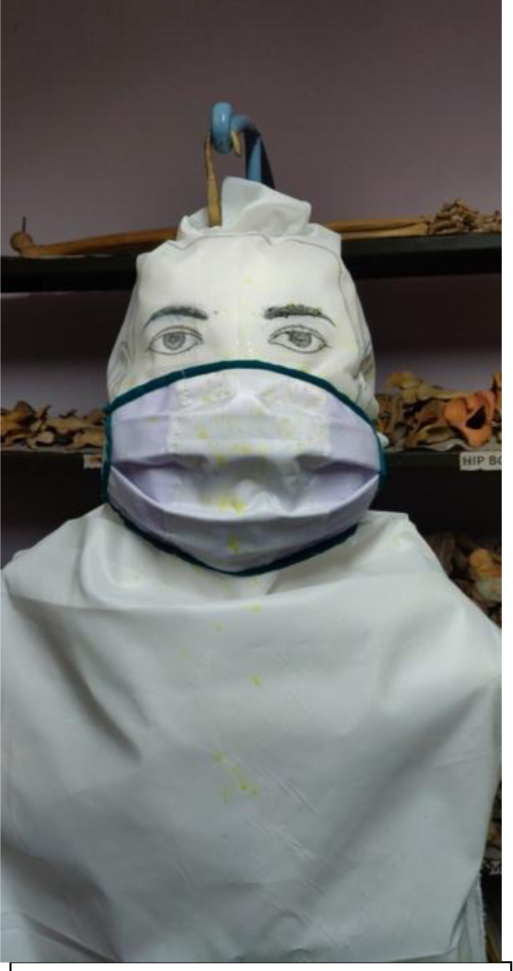
showing spread of aerosols/droplets on face and body under bright light

**Photo 14.**
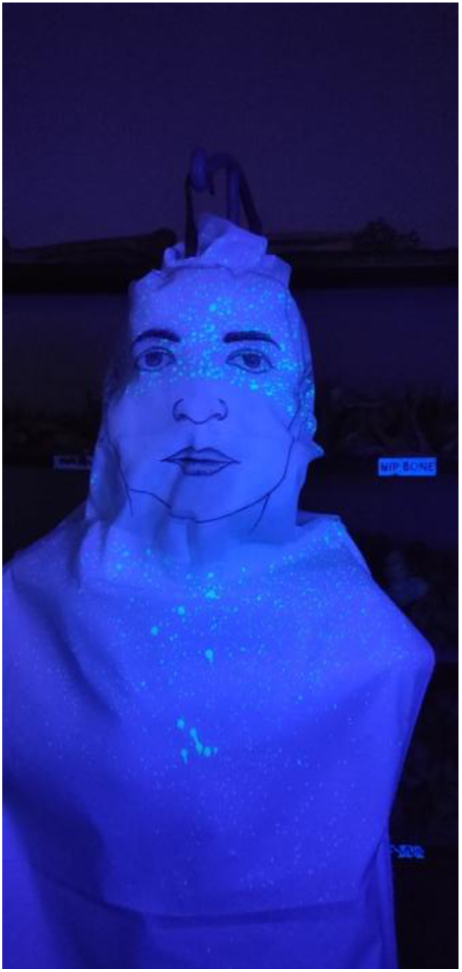
showing spread of aerosols/droplets on face and body after removal of mask under fluorescent light

## Discussion

This innovative mask was created by our laboratory after discussion with the end users of the general purpose masks from the community as well as health care workers. Main issues that derived from the discussion were- a) people were not sure of the efficacy of what they were wearing, b) the masks were either too big or too small, c) the masks with elastic strings were likely to be manipulated to cover only mouth or just the chin, d) reusability, e) price, f) role of face shield in the community, g) if cotton masks with some modifications could be used in hospital etc.

We tried to answer the above questions by creating this design, performing various experiments to test its efficacy and comparing it with other masks and N95FFR.

It was proven in our experiment that if people could maintain a distance of two meters, the chances of aerosols/droplets to reach their face are low. This chance increases if some turbulence is created either by fans or air-conditioner vents. In the open space the aerosols with viral particles will get diluted and may not cause infection; however, the same may not hold true for indoor spaces. Thus, wearing a mask indoors while other people are around should be a must. Also, even if others are not inside the public space it makes sense to keep the mask on to prevent contamination of surfaces.^6^

We created standard measurements for the mask by taking average of male and female faces in our area and these can be changed by others depending upon the region. We also put long strings to tie them behind the head to prevent the scope for manipulating the masks by the end user.

Since the inner two layers are of cotton, they can be washed any number of time. The outer impervious cloth has been found to be working for at least 40 reuses as per our previous experience.^5^

The cost for each mask was Indian rupees 50 (1USD= 76INR).

Community use of face shield has been discussed in previous studies^7^ and this worked in our experiment as well. Since, it was a manikin and was not breathing as human we cannot comment if aerosols would travel inside the face shield during inhalation.

We have observed that one of the two varieties of triple layers surgical mask was likely to be penetrated by the aerosols and droplets as it showed stain in the inner layer on microscopy. Since virus particles are smaller (120nm in diameter), there are likely chances that these gain entry to nose or mouth. Also, in cases where the wearer sneezes there will be even more chances that the aerosols/droplets will contaminate the surfaces while the wearer has false sense of safety. Use of impervious cloth in the middle half portion of the mask makes it safer. The impervious cloth can tolerate alcohol^5^ and can be sanitised with alcohol wipes for 30seconds, if exposure occurs.

The end users have liked the design and they did not find any breathing difficulty with half of the front layer being impervious.

To conclude, we feel that our innovation can be used in the community as well as general areas of the hospital like, offices, labs, etc. and can be a better alternative to single use triple layer surgical masks. Further testing may be done by other organizations to rule out bias in our study.

## Data Availability

If required, the raw data will be submitted by us.

## Acknowledgements

Miss Meenakshi Shahane, Mr Baba Jinde, Mr Ramesh Khajone, Mr Satish Shingare, Mr Ashok Wahiwatkar and Mr Shubham Dhok for their technical help.

